# Beyond olfactory cortex – severity of post-traumatic olfactory loss is associated with response to odors in frontal-parietal-insular networks

**DOI:** 10.1101/2020.06.09.20118539

**Authors:** Robert Pellegrino, Michael C. Farruggia, Dana M. Small, Maria G. Veldhuizen

## Abstract

Olfactory impairment after a traumatic impact to the head is associated with changes in olfactory cortex, including decreased gray matter density and decreased response to odors. Much less is known about the role of other cortical areas in olfactory impairment. We used fMRI in a sample of 63 participants, consisting of 25 with post-traumatic functional anosmia, 16 with post-traumatic hyposmia, and 22 healthy controls with normosmia to investigate whole brain response to odors. Similar neural responses were observed across the groups to odor versus odorless stimuli in the primary olfactory areas in piriform cortex, whereas response in the frontal operculum and anterior insula (fO/aI) increased with olfactory function (normosmia > hyposmia > functional anosmia). Unexpectedly, a negative association was observed between response and olfactory function in the mediodorsal thalamus (mdT), ventromedial prefrontal cortex (vmPFC) and posterior cingulate cortex (pCC). Finally, connectivity within a network consisting of vmPFC, fO, and pCC could be used to successfully classify participants as having functional anosmia or normosmia. We conclude that, at the neural level, olfactory impairment due to head trauma is best characterized by heightened responses and differential connectivity in higher-order areas beyond olfactory cortex.

**Significance Statement:** Olfactory impairment affects a quarter of the population, with subjective complaints usually confirmed with psychophysical measurements. Here, we demonstrate that the degree of olfactory impairment can also be categorized using neural responses to odors. Remarkably, regions with neural responses that were predictive usually showed an increase in response to odors with degree of impairment, rather than a reduction, as might be expected. Further, predictive cortical regions were not isolated to canonical olfactory areas.

---

A fifth (17 - 24%) of the population has an impaired sense of smell (1). Quantitatively the level of impairment may range from partial loss (hyposmia) to total loss (anosmia), while qualitative impairments include distortion (parosmia) or phantom smells (phantosmia). The etiology of these conditions may be peripheral, resulting from damage to the olfactory epithelium or its nerves, or central, with damage to either the olfactory bulb or primary / secondary cortices. Traumatic injury is among the main causes of olfactory impairment with 20 to 68% of impacts leading to olfactory function loss, depending on the severity of the trauma (2). Acute olfactory impairment is the most common occurrence after a traumatic impact with clinically significant recovery beginning around 3 months from the incident (3),(4). Yet, chronic impairment does occur regularly from trauma (5) and has a worse recovery rate than other etiologies (6),(7).

Olfactory impairment screening in the acute phase of a traumatic incident may constitute an important screening tool for long-term outcomes, as individuals with olfactory dysfunction have more anxiety and post-concussion symptoms a year from the time of trauma (1),(8). To objectively measure olfactory impairment an individual must complete a battery of tests (e.g.,Sniffin’ Sticks (9)); however, these tests are susceptible to inattention, cultural differences, and even deception. Recent advances in functional MRI (fMRI) analyses give rise to another option for predicting olfactory loss. Connectome-based Predictive Modeling (CPM), can be used to ‘fingerprint’ (or identify) an individual from a group based upon their brain connectivity profile (10). CPMs are able to predict fluid intelligence (10), attention (11), personality (12), adiposity (13), and drug abstinence (14). Distinguishing group differences in severity of olfactory impairment from fMRI has not been attempted but was suggested in a recent study (15). Additionally, to identify the neural correlates of post-traumatic olfactory loss may reveal mechanisms underlying loss and help identify treatment targets for rehabilitation and recovery.

To date, most studies of olfactory loss have focused on *structural* differences between healthy and impaired. Many of these studies include patients with different etiologies, including idiopathic origins. Here we briefly review only studies measuring acquired dysfunction, as congenital anosmia does not show equivalent changes in the brain (16). In general, grey matter volume decreases in primary (e.g. piriform) and secondary olfactory structures (e.g. insula, orbitofrontal cortex, anterior cingulate cortex and hippocampus) for patients with anosmia and atrophy increases with impairment duration (17),(18). The grey matter volume in the cerebellum, a brain area associated with the sensorimotor act of sniffing, also decreases (19). Similarly, a lack of olfactory afferent input decreases the volume of the olfactory bulb (20),(21) and the adjacent olfactory sulcus positioned between the gyrus rectus and medial orbitofrontal gyrus (22). The characterization of *functional* changes, whether found in voxel-wise activations or regional connectivity, has received far less attention than characterization of structural changes (23),(24). Spectral and olfactory event-related EEG discriminate among levels of severity in olfaction and correlate with the amount of loss (25),(26),(27),(28). However, an absent signal from scalp electrodes does not guarantee the inability to smell (29), although recent advances with olfactory bulb EEG indicate improved specificity (30). Previous functional MRI (fMRI) studies often are difficult to interpret because of methodological issues, including small sample sizes (31),(32),(33), no control group (34), or the use of bimodal odors that induce both olfactory and trigeminal responses (35),(36),(37),(38). However, a handful of rigorous studies do indeed exist and these suggest that olfactory loss is reflected in decreased response in olfactory circuits that are often associated with loss duration. For example, a recent study reported activation of the right primary olfactory cortex was graded according to olfactory ability with a response greater in hyposmia than anosmia and greater in normosmia than hyposmia. Loss duration did not influence this pattern, but in the contralateral (left) primary olfactory cortex response was negatively associated with the duration of impairment for patients with hyposmia but not patients with anosmia, while response in the left insula was negatively associated with duration of loss in patients with anosmia (39), a finding that has been replicated (40),(24). Studies have also demonstrated reduced anterior cingulate (41) and right orbitofrontal cortex activation in hyposmia (39), (41) and anosmia (39). Rather than decreased responses, functional impairment may also be characterized by increased responses to odors vs odorless. To this point, increases in activation to odors have been demonstrated in parahippocampal and parietal cingulate cortex (40),(41), as well as connnectivity in a functional network consisting of olfactory, somatosensory and integration regions in response to bimodal odors (38).

The aim of the current study was to test whether the neural response to odors can be used to discriminate between patients in different categories of olfactory impairment severity. Additionally, we wanted to identify the neural correlates of post-traumatic olfactory loss. To achieve these objectives, we used fMRI to measure the neural responses to odors compared to an odorless control in a relatively large post-traumatic patient population with a range of olfactory impairment; hyposmia and functional anosmia, compared to a similarly sized healthy control group. Control and patient groups were analyzed with voxel-wise analysis and CPM for a predictive model. We hypothesize that reduced responses in canonical olfactory cortex (piriform, insula and orbitofrontal cortex) will be associated with olfactory impairment and these networks have predictive value.

## Methods

### Participants

Patients who entered hospital treatment for traumatic impact were evaluated for study eligibility. These patients underwent a standard ear, nose, and throat (ENT) examination with an endoscope as well as objective olfactory testing with Sniffin’ Sticks. Patients having an age-adjusted composite score within the hyposmia (∼ 16 – 32) or functional anosmia range (< 16) were invited to participate in the study (9). “Functional” anosmia is a quantitatively reduced olfaction to the extent that a subject has no function useful in daily life while anosmia is the absence of all olfactory function (1). In this report, the “anosmia” group includes individuals with “functional” anosmia as well as total olfactory loss or “true” anosmia, so it should be noted that there may still be residual function in individuals in this group. We will refer to this group from here on as “anosmia”. All participants filled out a medical questionnaire and had no major comorbidities, acute diseases, or took medicine that would significantly impact their sense of smell. A control sample (n=22) was recruited in and around the campus of the Technical University and hospital in Dresden. Participants in the control sample were tested, and scored within the healthy olfactory functionality range (∼ > 32) and will be described as the “normosmia” group hereafter. Participants with normosmia reported that they had no previous head traumas nor olfactory issues while the patient groups reported no olfactory issues prior to their trauma. Traumatic damage severity scores were calculated for a subset (50 out of 63) of the patients (by a neurologist on staff that inspected the anatomical MRI images) with a range from none to severe depending on the 11 brain regions evaluated (42). This quantification is acomposite of both degree of lesion and the number of brain regions affected. We also calculated a sum score of regions affected regardless of degree of lesion and refer to that as “extent scores). Previous research has shown the severity of damage correlates with olfactory function (18). We evaluated the severity and extent scores in our subset of 16 out of 22 participants with normosmia, 15 out 16 participants with hyposmia and 19 out of 25 participants with anosmia and observed that the groups differed on these scores, such that there was for more widespread and severe damage in participants with anosmia/hyposmia relative to participants with normosmia. This same numerical trend was present for the difference between the anosmia and hyposmia groups, however the effects here did not reach significance (see Supplementary Table 1 and Supplementary Fig 1).These observations confirm that olfactory dysfunction here reflects degree of brain trauma. Participants were informed of the details of the study and signed a consent form approved by the Ethics Committee of Medical Faculty Carl Gustav Carus at the Technical University of Dresden in accordance with the Declaration of Helsinki.

## fMRI Procedure and Processing

### Procedure

For fMRI acquisition, participants underwent a block design during which the common, culturally-relevant odors peach and coffee were delivered at neat concentrations (peach and coffee; Frey & Lau, Henstedt-Ulzburg, Germany). These odors were selected from pilot studies with an expert panel (n=6) and produced little or no trigeminal sensation as indicated by ratings. Odors were presented intranasally with an olfactometer (2 L/min flow) to each nostril for 9 6 scans across alternating ON and OFF blocks (starting with OFF which presented humidified a ir), for four sessions of blocks. The ON blocks were 20 seconds long with stimulus a duration of 2 seconds followed by 1 second interstimulus intervals (ISIs). There were no differences between ON and OFF blocks besides one had an odor in the airstream and the other did not. At the end of each of the four conditions, individuals were asked to verbally rate the intensity [not perceived (0) to extremely strong (10)] and valence [very unpleasant (−5) to very pleasant (5)] of the odor as well as try to freely identify the odor (4 odors, expressed as proportion correct). Participants had the study design explained to them and were asked to breath normally throughout the scanning session. Each session took approximately 45 minutes.

### fMRI Scanning Parameters

A 1.5 T magnetic resonance imaging scanner (Siemens Sonata; Siemens, Erlangen, Germany) with a full-head eight channel receiver coil was used for image acquisition. A gradient echo T2-sensitive echo planar imaging sequence was employed for 96 functional volumes/conditions in thirty-three slice locations, covering the entire head (repetition time [TR]: 2500 ms, echo time [TE]: 40 ms, image matrix: 64 × 64, in-plane resolution: 3 mm, through-plane resolution: 3.75 mm). Our TE was selected because it had been established for 1.5 Tesla scanners for the imaging of limbic structures (43). Images were acquired in the axial plane oriented parallel to the planum sphenoidale to minimize artifacts. A full brain (192 slices) T1-weighted turbo FLASH three-dimensional sequence was acquired to overlay functional data (TR: 2,180 ms, TE: 3.93 ms, slice thickness: 1 mm).

## Data Analysis

### Demographic and behavioral data

Demographic and behavioral data were analyzed with GraphPad Prism 7.01 (GraphPad Software Inc.). We calculated normality for impairment duration, TDI, the TDI subscales, age, free ID, intensity and pleasantness ratings. For all variables except impairment duration, we used an ordinary one-way ANOVA with group as the between-subjects factor if normality was not violated, and Kruskal-Wallis ANOVA if normality was violated for one or more groups. To test for differences in impairment duration between hyposmia and anosmia, we used a non-parametric Mann-Whitney U test. Post-hoc t-tests to assess differences between each of the groups were corrected for multiple comparisons using a two state linear step-up procedure of Benjamini, Krieger and Yekuteli for ordinary ANOVA and Dunn’s procedure for Kruskal-Wallis ANOVA. For gender distribution differences across groups we performed a Chi-square test. Correlations were calculated with Pearson’s coefficient. All analyses used an alpha of 0.05 to determine significance.

### MRI Preprocessing

Data were analyzed on Linux workstations using MATLAB R2011a (MathWorks) and SPM12 (Wellcome Trust Centre for Neuroimaging, London, UK). Functional images were realigned and coregistered to the T1 image. The anatomical T1 image was processed using a unified segmentation procedure combining segmentation, bias correction, and spatial normalization (44). The same normalization parameters were then used to normalize the functional images. All functional images were detrended using a method for removing any linear components matching the global signal at each voxel (45). Finally, functional images were smoothed with a 6 mm FWHM isotropic Gaussian kernel.

### Voxelwise MRI analysis for group differences

In a voxel-wise analysis aimed at isolating brain regions that respond to odor vs odorless in all olfactory impairment groups. For the time-series analysis on all participants’ data, a high-pass filter (300 s) was included in the filtering matrix (adjusted f rom the convention in SPM12 to reflect the longer period between two blocks) to remove low-frequency noise and slow drifts in the signal. Condition-specific effects at each voxel were estimated using the general linear model. The response to events was modeled by a canonical hemodynamic response function included in SPM12. The temporal derivative of the hemodynamic response function was also included as part of the basis set to account for up to 1 s shifts in timing of the events (46). There were two events of interest, “odor” and “odorless”. For “odor”, all odor ON blocks (regardless of nostril or odor quality) were collapsed into a single event. For “odorless” we modeled all OFF blocks. Each event had a duration of 20 s. The Artifact Detection Tools (ART) toolbox for MATLAB was used to detect global mean and motion outliers in the functional data (Gabrieli Laboratory, McGovern Institute for Brain Research, Cambridge, MA, USA). Motion parameters were included as regressors in the design matrix at the single-subject level. In addition, image volumes in which the z-normalized global brain activation exceeded 3 SDs from the mean of the run or showed 1 mm of composite (linear plus rotational) movement were flagged as outliers and weighted during SPM estimation. For each participant we created a single contrast of interest: odor minus odorless. To assess the effect of olfactory impairment we created a second-level analysis with between-subjects factor “group.” The parameter estimate images of odor vs. odorless for each participant were entered into a one-way ANOVA. We included covariates-of-no-interest for age and gender. We created a T-contrast for the average effect of group and a linear F-contrast of group. The t-map threshold was set at puncorrected < 0.005 and a minimum 5 voxel cluster size. Clusters were considered significant at P < 0.05 Family Wise Error (FWE) corrected at the cluster level. A less stringent region of interest (ROI) approach was used for responses in predicted regions of olfactory cortex. We performed small volume searches using spheres (6 mm radius) around coordinates in thalamus, insula, and piriform from previous work (47).

### Group MRI equivalence testing

We used Han et al. to determine an expected meaningful effect size for differences between groups in piriform cortex (39). We then tested for the significant absence of a difference (or the equivalence) of the functional anosmia and normosmia groups with the “Two One-Sided Tests” (TOST) procedure (48),(49). This procedure can be used to determine if an effect size is surprisingly small compared to an existing effect. We used the TOST two-sample student t-test with parameter estimates extracted from the peak voxel in the four clusters in piriform cortex (Supplementary Table 2) where we observed a main effect of odor-odorless.

### Functional MRI connectivity and Predictive Modeling

We performed Connectome-based Predictive Modeling (CPM) on the entire time-series acquired to examine whether whole-brain functional connectivity correlates with olfactory impairment category (i.e. functional anosmia, hyposmia, or normosmia). Normalized, motion-corrected images were preprocessed in BioImage Suite for use in our functional connectivity analysis **???** correction at this stage included regression of 24 parameters of motion, comprising six rigid-body motion parameters, their temporal derivatives, and their squares (50). We regressed the mean time courses of the global signal, CSF, and white matter from the data, implemented linear trend removal and low-pass filtering. We then created functional connectivity matrices for each participant using a 268-node whole-brain parcellation (51). Functional connectivity matrices were created in BioImage Suite by averaging the BOLD signal among all voxels within a given node and correlating, using Pearson’s r, this time course with those obtained from every other node. This process was then repeated iteratively until a 268×268 correlation matrix was obtained for each participant. Connectivity matrices were then Fisher Transformed to convert the skewed distribution of r values to an approximately normal distribution. Matrices were averaged across runs to generate a mean matrix per participant, which was then collapsed across participants to yield a 268×268xN-participant matrix.

CPM implements linear regression to correlate, using Pear-son’s r, each edge (i.e. connection) in each connectivity matrix with TDI scores or ‘impairment group membership’ per participant. Because CPM is not optimized to discriminate among multiple classes, we used it as a binary classifier to discriminate between functional anosmia and normosmia, normosmia and hyposmia, and functional anosmia and hyposmia patient statuses. In this case, status was coded as either a 0 or 1 and estimates for these values were rounded either up (to 1) or down (to 0) to determine model accuracy. Following correlation, each edge is subsequently associated with a p-value and a threshold is applied, in this case p = 0.01, to determine the most relevant edges to build our brain-behavior model. Two networks are then created, a positive network comprised of positive edges (i.e. positively correlated with behavior), and a negative network comprised of negative edges (i.e. negatively correlated with behavior). A single subject summary value, ‘network strength’ is then calculated for both positive and negative networks by summing their respective strengths. This analysis uses a leave-one-out cross validation framework, with models created on N-1 participants to relate positive and negative network strengths to behavior. The model is then applied prospectively to the left-out participant’s network strengths to generate an estimated behavioral score.

Correlations between observed and estimated scores were only obtained for TDI scores. To determine the significance of these correlations, we conducted permutation testing; here, 1000 repetitions of CPM with randomly shuffled observed scores were used to generate estimated scores and networks. The 1000 correlation coefficients comprised a null distribution against which the correlation coefficients obtained in results were tested for significance. The final p-values were the number of permutations out of 1000 that resulted in higher correlation coefficients than those reported in results.

Average frame-to-frame displacement (FTF) was used as a covariate in our CPM analysis due to the confounding nature of subject motion in functional connectivity analyses. In brief, FTF was calculated by taking the Euclidean distance from the center of gravity of one image to the next, summing these distances within a run, and then averaging across runs to create a single score per participant (50).

## Results

### Participant characteristics

Participants consisted of 63 individuals with either clinically defined normosmia, hyposmia, or anosmia. On average, participants were 54.4 years of age (Std 13.2, range 22-75). Overall, the effect of group on age was marginally significant (p = 0.091, Supplementary Table 1), but post hoc t-tests (corrected for multiple comparisons) showed participants with anosmia were significantly older than their counterparts with normosmia (p < 0.05, Fig 1). In total more men participated (25 women, 38 men); however, there were more women (10) than men (6) with hyposmia (Fig 1, with a marginally significant effect of group on gender (p = 0.097, Supplementary Table 1). Therefore, we included age and gender as covariates-of-no-interest in the subsequent fMRI analyses. Duration of impairment ranged from 2.03 to 211 months and was not significantly different between anosmia and hyposmia groups (Fig 1, Supplementary Table 1). Threshold, discrimination, identification and composite TDI scores at time of clinical testing showed significant differences in all olfactory domains of the Sniffin’ Sticks between all groups (p < 0.05, Fig 2, Supplementary Table 1).

**Fig. 1.**
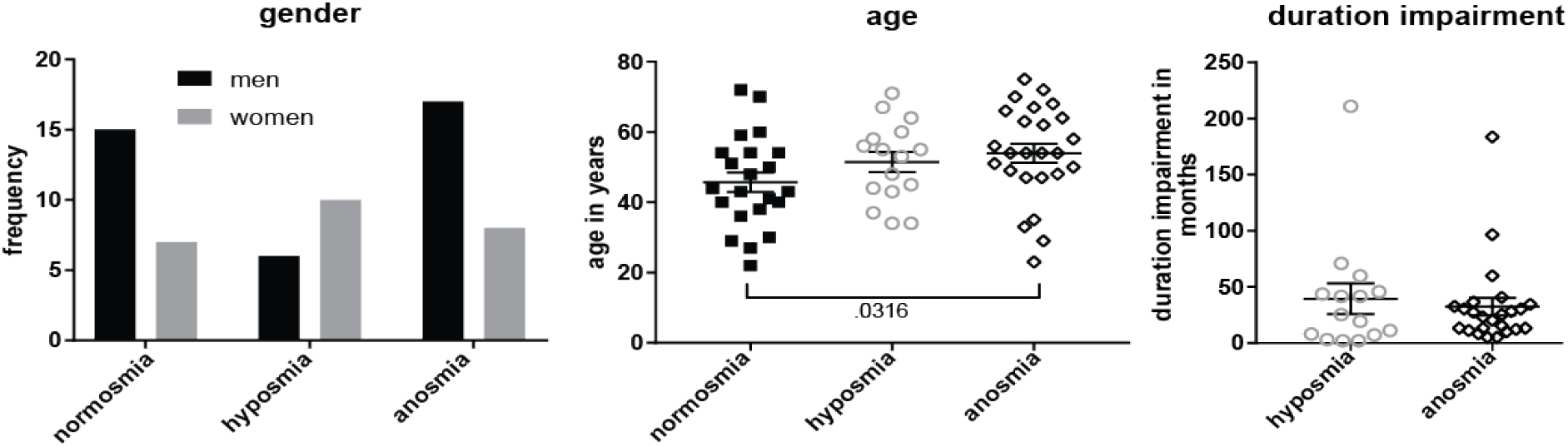
Participant gender and age by group. Left panel shows a histogram of men (solid black bars) and women (solid gray bars) count. Center panel shows mean (center line) age with standard error of the mean bars (shorter upper and lower bars), with individual data points overlaid for normosmia (solid black squares), hyposmia (open gray circles), and functional anosmia (open black diamonds) participant groups. Right panel shows the duration of olfactory impairment in months, showing the relevant hyposmia and anosmia groups only. Bracket indicates significant post-hoc t-test between groups (corrected for multiple comparisons) with p-value.

**Fig. 2.**
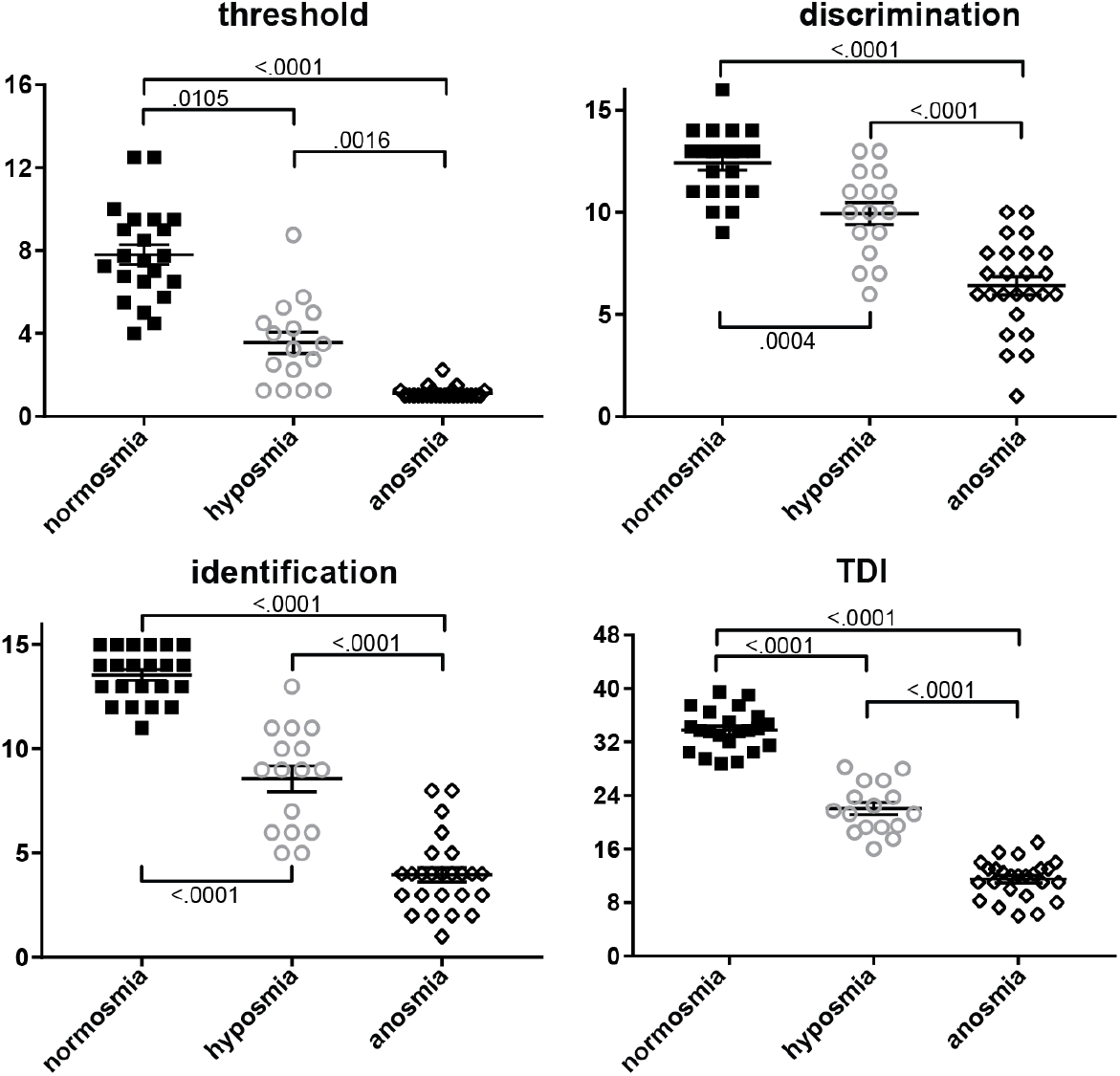
Threshold (upper left panel), discrimination (upper right), identification (lower left), and composite TDI (lower right) score from Sniffin’ Sticks test by group. Legend details as in Fig 1.

### Olfactory function at fMRI scan

The odors presented in the fMRI scan session were rated as less intense by participants with anosmia or hyposmia than by participants with normosmia, and participants with anosmia in turn rated the odors as less intense than participants with hyposmia (Fig 3). Ratings of pleasantness of the odors displayed a weaker relation with olfactory impairment group. Participants with anosmia rated the odors as less pleasant than participants with normosmia or hyposmia, but there was no significant difference in pleasantness ratings between the participants groups with normosmia and hyposmia. Free identification (no response categories provided) of the odors was significantly impaired in participants with anosmia, with most participants giving incorrect responses only, compared to the participants with hyposmia and normosmia. Most participants with normosmia gave 100% correct responses on free identification, and performed significantly better than the participants with hyposmia. Note that while performance on free identification of odors follows the general pattern of olfactory impairment, these scores also show a substantial amount of variation; some participants with hyposmia identify 100% of the odors correctly, while others with hyposmia identify 0% of the odors correctly. Likewise, we observe a large overlap in pleasantness ratings of the odors across groups. TDI correlates well with the olfactory function measures obtained at the fMRI scan (all r’s > .73, except pleasantness ratings; Fig 4), indicating that neural response to the odors used in the fMRI scan is representative of olfactory impairment as assessed by Sniffin’ Sticks. Pleasantness of the odors is the only variable that shows little to no relation with the other variables.

**Fig. 3.**
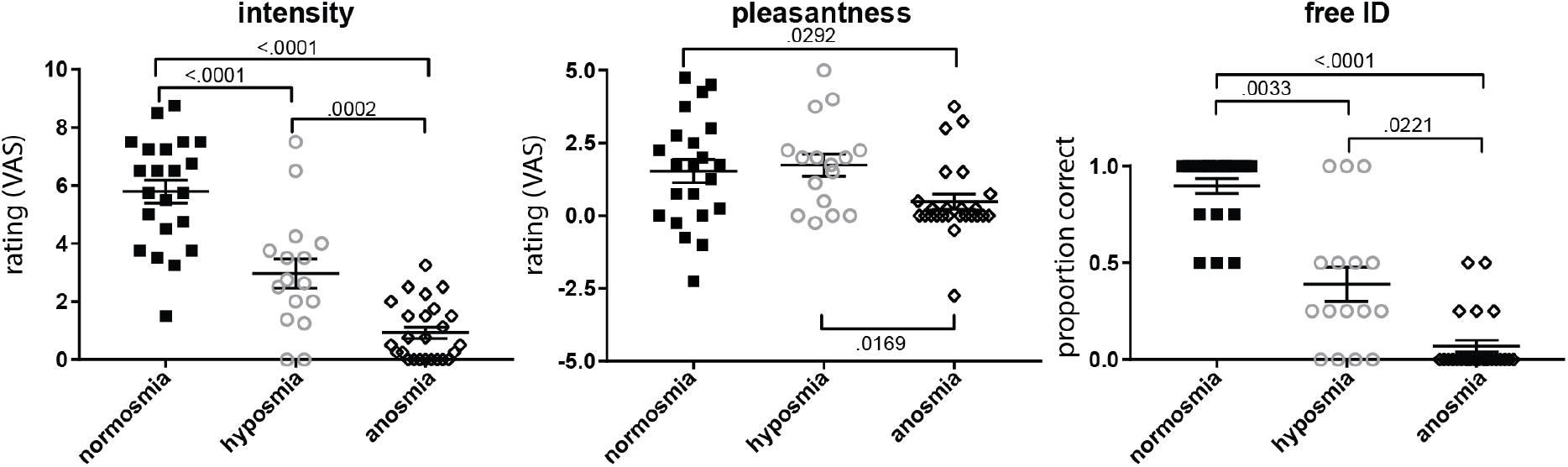
Perceived intensity (left panel), pleasantness (center), and free identification (right) of odors by group. Legend details as in Fig 1

**Fig. 4.**
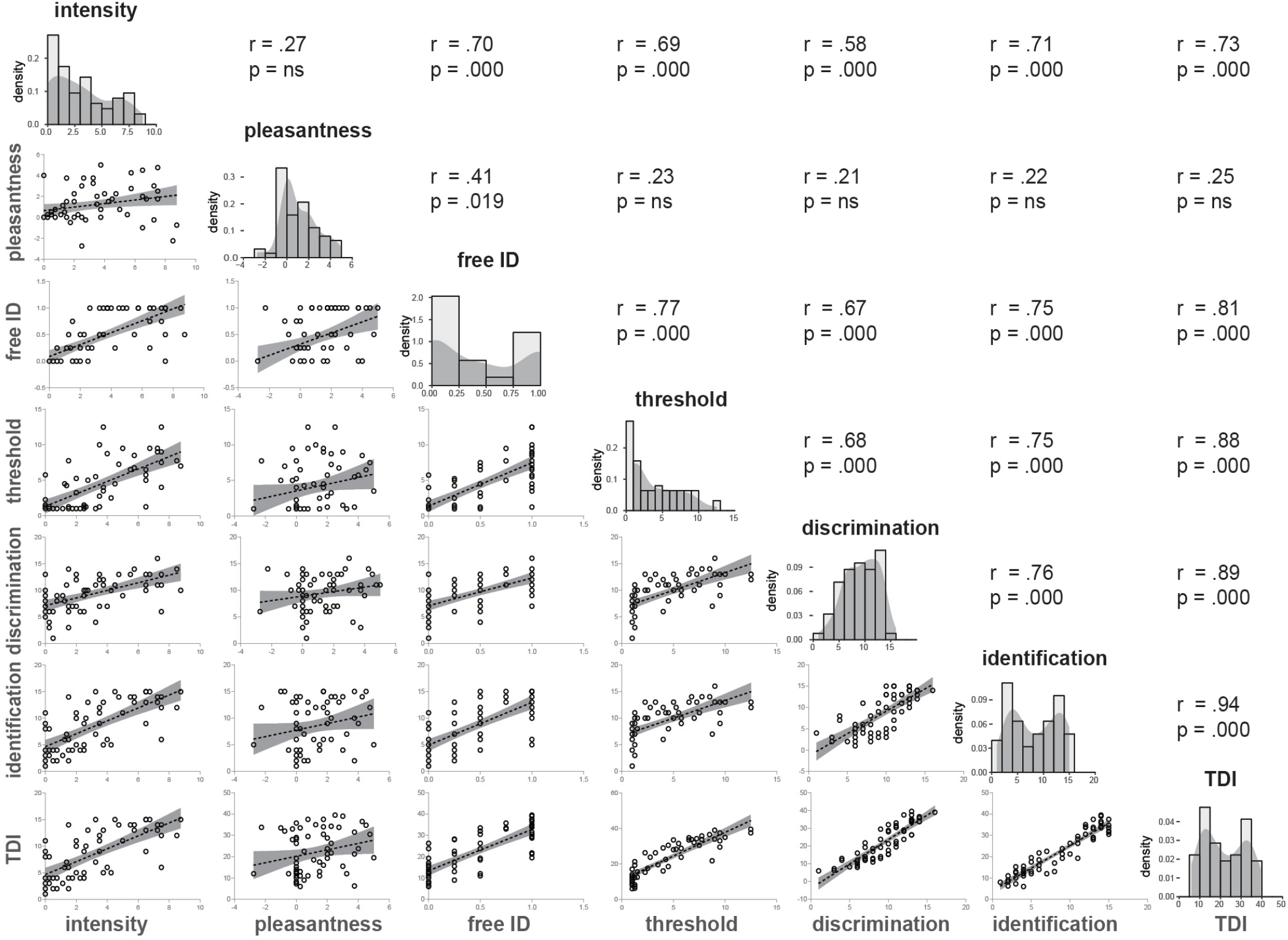
Cross-correlations between olfactory function measures and distribution of scores regardless of group. Diagonal shows histograms and density plots of distribution of scores. Below the diagonal scatterplots illustrate relation between variables indicated at the bottom of each column and the left of each row. Lines illustrate best fit of linear regression, with transparent gray area indicating 95Above the diagonal the Pearson correlation and p-value of significant statistic are given. ns = not significant.

### Voxel-wise neural response to odor vs odorless for all olfactory impairment groups

In a voxel-wise analysis aimed at isolating brain regions that respond to odor vs odorless in all olfactory impairment groups, we examined a T-contrast for the average effect of group. Each group exhibited a positive response in right anterior piriform (aPir)/ventral insula (Fig 5A and Supplementary Table 2). A similar pattern of results was observed for left aPir and bilateral posterior piriform (pPir). While these latter peaks were not significant for multiple comparisons, we report them in Fig 5A and Supplementary Table 2. This serves to illustrate a similar magnitude of response across groups in all areas of piriform cortex, including those areas that others previously showed to differ between normosmia and anosmia (39). Each group shows an average neural response to odors vs odorless stimuli. (i.e. a parameter estimate above 0). While in each group there may be individuals that showed a smaller response to odor compared to odorless (i.e. a parameter estimate below 0), such individuals were present in each group in similar proportions relative to the total group size. Equivalence tests confirm the absence of a difference between the anosmia and normosmia groups in the clusters in piriform cortex (Supplementary Table 3). We additionally observed unpredicted responses to odor vs odorless across all groups in intra-parietal sulcus (iPS), cerebellum and inferior frontal gyrus (iFG) (Fig 5B and Supplementary Table 2).

**Fig. 5.**
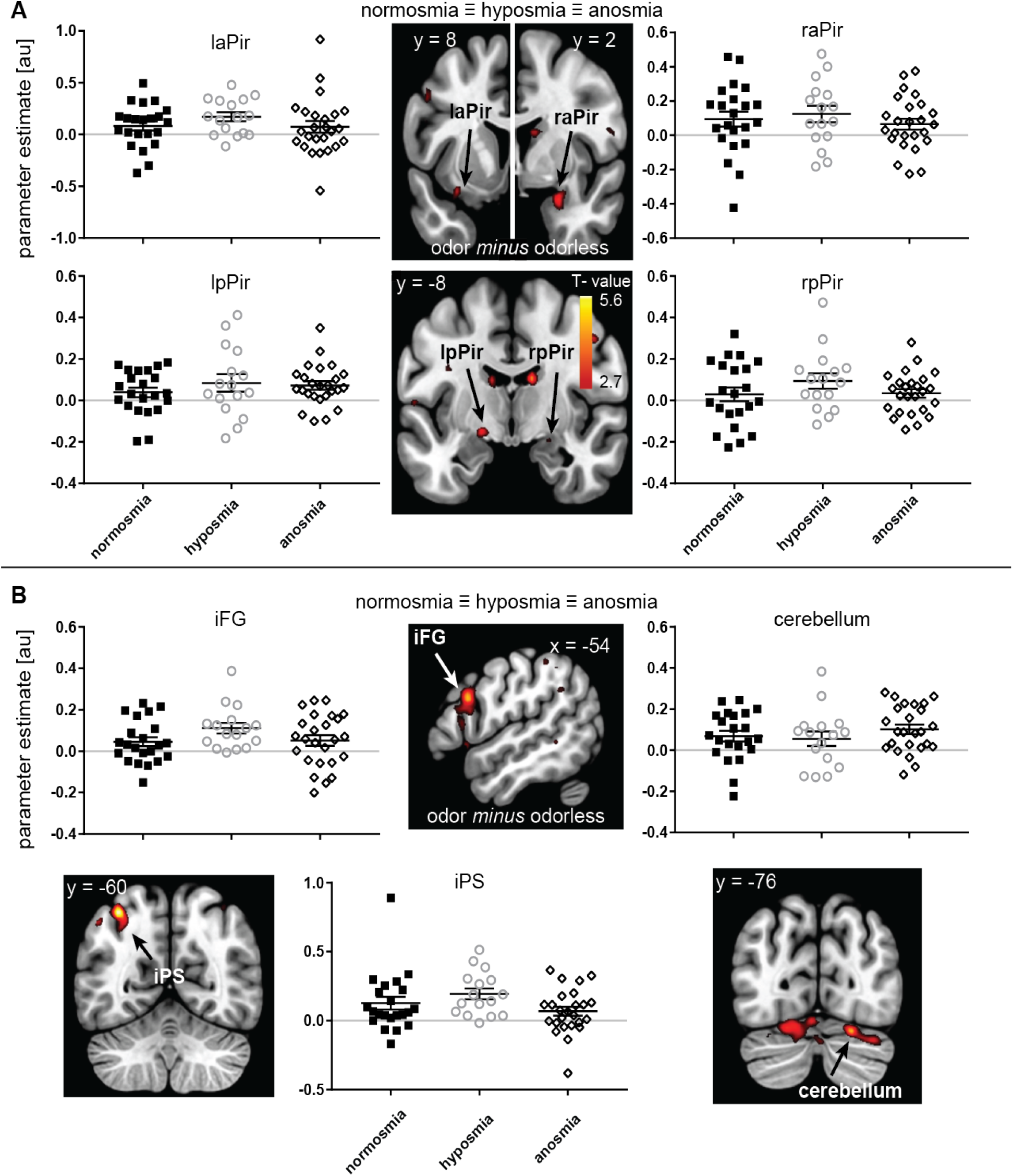
Neural response to odor-odorless regardless of olfactory impairment, A. in ROI of piriform cortex, and B. in unpredicted regions. Sections (slice location indicated in MNI-coordinate) show canonical anatomical template with SPM T-map overlaid, thresholded at puncorrected <.005, and a minimum of 5 contiguous voxels. Color gradient scale depicts supra-threshold T-values. Graphs show parameter estimate (in arbitrary units) for the voxel with the peak t-value in the cluster on the y-axis. For illustrative purposes we plotted the a line through y = 0, to illustrate that, on average, each group shows positive neural response to odor vs odorless in anterior and posterior piriform (aPir and pPir), inferior frontal gyrus (iFG), intra parietal sulcus (iPS) and cerebellum. Legend details as in Fig 1

### Voxel-wise neural response to odor vs odorless as a function of olfactory impairment

Next, in an additional voxel-wise analysis, we specified an F-contrast of linear changes in neural response as a function of group to isolate brain regions that respond in a pattern of increasing neural response with increased olfactory impairment (positive linear trend) or increased olfactory function (negative linear trend). We observed multiple clusters in pCC, vmPFC, and bilateral mdT in which participants with anosmia had a stronger response than those with hyposmia and those with hyposmia in turn had greater response than those with normosmia (Fig 6A and Supplementary Table 4). Conversely, we observed greater response in participants with normosmia than those with hyposmia and greater response in those with hyposmia than those with anosmia in the left aI/fO and right fO (Fig 6B and Supplementary Table 4).

**Fig. 6.**
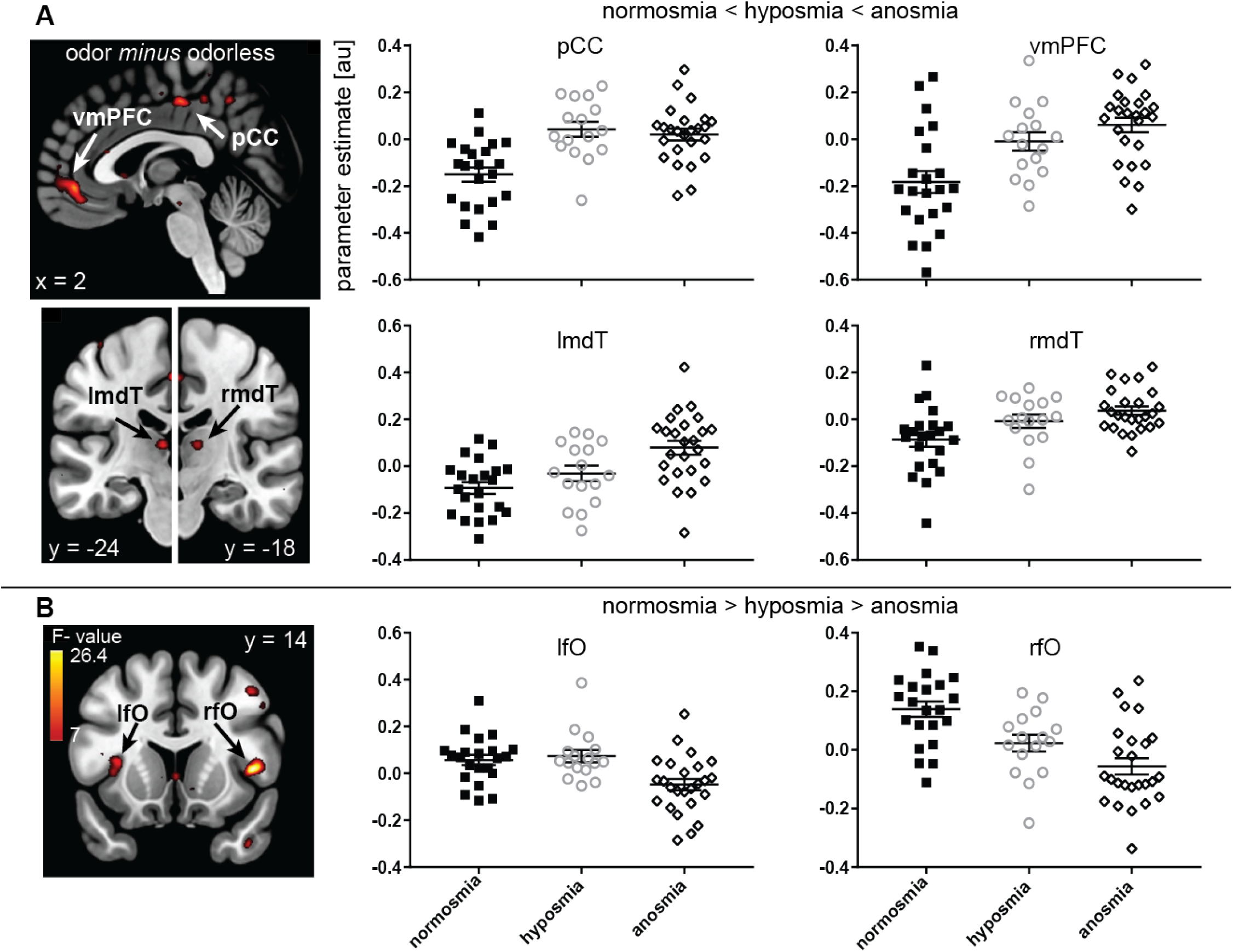
Neural response to odor minus odorless as a function of olfactory impairment. A. clusters in posterior cingulate cortex (pCC), ventromedial prefrontal cortex (vmPFC) and bilateral medio-dorsal thalamus (mdT) showing increasing neural response with increased olfactory impairment, such that participants with anosmia have stronger response than those with hyposmia and those with hyposmia in turn have greater response than those with normosmia. B. clusters in bilateral frontal operculum and left anterior insula (lfO/rfO) showing increased neural responses with increased olfactory function, such that participants with normosmia have greater response than those with hyposmia and those with hyposmia in turn have greater response than those with anosmia. Color gradient scale depicts supra-threshold F-values (thresholded at p uncorrected <.005). Legend details as in Figs 1 and 5

### Functional connectivity and group classification

Using a leave-one-out cross validation framework, we determined a whole-brain functional connectivity network that explains approximately 17% of the variance in TDI score in a novel individual after controlling for age, which we know from the literature and from this cohort (p = 0.02, r2 = 0.078) to negatively predict TDI (52). While this model was significant (p < 0.001), we noted a very large mean squared error (110.93), which hinders model interpretation and utility. Therefore, we undertook a binary classification approach, aiming to use CPM to determine a connectivity network capable of discriminating between two of the groups (i.e. between anosmia and normosmia, between anosmia and hyposmia, and between hyposmia and normosmia). We found that this network can discriminate between individuals with anosmia or normosmia with a combined accuracy of approximately 64% after controlling for age. That is, in 64% of cases, an individual was correctly classified as either having anosmia or normosmia. The sensitivity for detecting anosmia in this sample is 72%, while the specificity is 55%. The positive predictive value (PPV) for this test was 64%, while the negative predictive value (NPV) was 63%. Notably, controlling for motion, gender, or impairment duration did not appreciably alter these results. This network contains highly connected nodes that correspond to regions from our voxel-wise analysis (i.e. Fig 6A-B) such as right aI/fO, vmPFC, and pCC (Fig 7). We furthermore found high-degree nodes in regions outside of vmPFC and pCC, such as the striatum and midbrain, However, we did not see mdT emerge among the 10% high-degree nodes (Supplementary Table 5); though this does not necessarily imply that it does not play a role in classification. This method could not reliably distinguish between the other groups; hyposmia and normosmia, or hyposmia and anosmia.

**Fig. 7.**
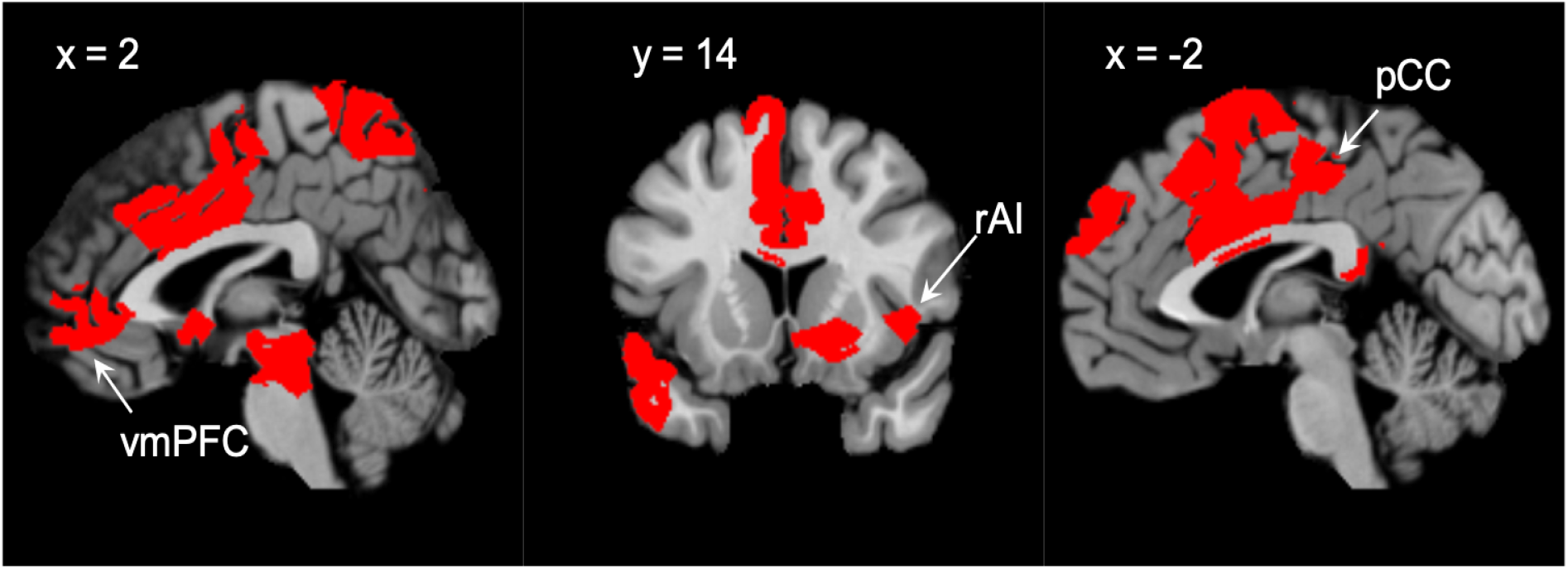
Nodes, or regions, with the highest number of connections in the brain-behavior model include members of both default mode (vmPFC and pCC) and salience (rfO/raI) networks.Threshold for activation clusters at p <.01.

## Discussion

The goal of the current work was to test whether neural response to odors could be used to discriminate between patients in different categories of olfactory impairment severity. We predicted that reduced responses to odors vs odorless in the canonical olfactory cortex (piriform, insula and orbitofrontal cortex) would be associated with olfactory function. As predicted, olfactory loss was associated with reduced responses in anterior insula/frontal operculum. However, we did not observe reduced responses in other canonical olfactory areas, and we unexpectedly observed increased responses in mdT, pCC, and vmPFC. Moreover, a functional connectivity network that contained several of the neural correlates that differed between groups in our voxel-wise fMRI analyses – fO, pCC, and vmPFC, was able to (within-sample) predict olfactory function differences between anosmia and normosmia at above-chance levels. To our knowledge, this is the first model capable of categorizing those with and without smell at above-chance levels using functional brain scans.

The insula and overlying frontal operculum are known to play a role in chemosensation, with olfactory projections terminating in the ventral and posterior agranular insula (53) and indirect connections to frontal operculum through orbitofrontal cortex (54). The region isolated here was dorsal to the agranular insula areas. This region shows consistent activation in odor vs. non-odor comparisons across studies (55) and a recent parcellation of primary olfactory areas in humans shows that frontal operculum and anterior insula are both functionally connected with piriform cortex (56). In normosmia, this region of the insula and overlying operculum is involved in attention to odors (47) (55), and may reflect awareness of odors or engagement in any task related to odors. In contrast, pCC and vmPFC responses to odors vs odorless were enhanced in those with smell loss. These two regions are often reported in a network of regions known as “default mode network”, that engages during times of rest or the absence of a task (57). The specific pattern of decreased and increased responses to odors with olfactory impairment observed here suggests that patients with functional anosmia may be displaying lapses in task-engagement (58). This may be due to the lack of input from an odor-related task; or put another way, the patient is in a state of boredom during a long scanning session where nothing happens and and their “mind wanders” (59). However, it is important to note that patients with hyposmia, who typically can still smell something, showed roughly similar responses to the functional anosmia group in the pCC for example (see Fig 6). In contrast, the hyposmia group showed intermediate responses (between functional anosmia and normosmia) in the vmPFC. This suggests that there may be more than one process reflected in the differences between groups we observed. Indeed, it has been noted that the default mode network may reflect mulitple interwoven networks (60). Intermediate responses in hyposmia would be consistent with other suggested functions of the default network such as memory retrieval (60) a type of processing previously speculated in this patient population (41). Another possibility is that activations in pCC and vmPFC reflect (negative) self-referential thinking, with patients with decreasing function experiencing greater degrees of frustration regarding their inability to perceive the odors presented to them (61). Lastly, participants were asked to make a variety of judgements in during brief breaks between blocks (no scanning) related to odor intensity and quality and reflections on those judgements may be affect neural response during scanning. Future studies may explicitly test the role of these areas in olfactory (dys)function and their potential (mal)adaptive role in olfactory recovery, rehabilitation and/or training.

Our voxel-wise MR analyses showed an increase in mdT response to odors vs odorless with impairment. Response in mdT has been shown to be associated with attention to and encoding of odor stimuli (62),(63), as well as odor novelty or task complexity (64). Perhaps increased mdT response in the current work reflects increased direction of attentional resources towards processing of signals in primary olfactory cortex that are not advancing to regular higher-order processing and/or attempts to recognize odors. Beyond its specific role in olfaction, mdT has broad connectivity to the pCC and vmPFC regions and it has been suggested that pulivnar and mdT may be involved in suppression of task positive networks and/or enhancement of the default mode network (60). Thus it is also possible that mdT responses here reflect its mediating role in focusing on internal processes, such as sensory memories. However, we did not find evidence for a role for the mdT in the connectivity network that can classify participants in the normosmia and functional anosmia groups. This apparent contradiction likely reflects the inherent differences in metrics used (i.e. functional connectivity versus voxel-wise responses); we used both approaches for enhanced rigor and because they each uniquely contribute information about how olfactory impairment affects neural responses. CPM relies on a 268-node functional parcellation of the brain, rather than an anatomical parcellation, which could obscure effects from small nuclei in the thalamus.

We observed no difference in piriform activation across olfactory impaired groups and their healthy counterparts as well as no evidence of piriform cortex connectivity contributing to our model. Most studies in patients with olfactory loss do not observe differences in piriform cortex responses to odors (40),(41),(24), however, to our knowledge we are the first to explicitly assess whether responses in piriform cortex are of similar magnitude across functionally different groups with reference to an expected effect size from another study (39). Why would patients with olfactory loss still show a neural response to odor vs odorless in primary olfactory cortex? This may be understood in the context of the type of olfactory impairment, in this case post-traumatic, which may lead to functional anosmia, but not complete anosmia (as observed in congenital cases for example). Reichert et al. (24) demonstrated piriform activation in response to sniffing clean air among patients with functional anosmia, and suggested that these patients may still have partially intact pathways from the olfactory epithelium to the piriform, but no higher-order processing leading to perception. Such patients may not be aware of odors, but still show neural responses to odors. This is consistent with numerous studies showing that unconscious odor detection may alter brain activity (65),(66),(67), and even behavior (68). Functional neuroimaging studies on patients with isolated congenital olfactory impairment, which to our knowledge do not exist at this time, may lend substance to this explanation. Interestingly, very recently a report on intact resting-state networks in olfactory areas were confirmed in congenital anosmia (69). The current results suggest that the post-traumatic damage causing olfactory dysfunction is central and in other brain areas than the primary olfactory cortex, however, it is also possible that this network reflects regions that have adapted to cope with olfactory loss.

## Conclusion

With increased post-traumatic olfactory function (normosmia > hyposmia > functional anosmia) we observed greater responses to odor vs odorless stimuli in anterior insula, but not in piriform cortex. In addition, we observed reduced responses with increased olfactory function in mediodorsal thalamus, ventromedial prefrontal cortex and posterior cingulate cortex. Connectivity in a large-scale network that includes anterior insula, ventromedial prefrontal cortex and posterior cingulate cortex discrimates between patients with anosmia or normosmia. These results imply that olfactory function in the central nervous system is not best captured by responses to odor vs odorless in canonical olfactory cortex. Rather, olfactory function is best characterized by connectivity in functional networks excluding canonical olfactory cortex. Future studies should focus on testing the role of these functional networks and whether modulation of these networks may improve function.

As hinted at in past papers, objective diagnosis of olfactory impairment is possible with functional scans via fMRI implicating its potential clinical usefulness. Similar to EEG (70), this neuroimaging technique provides a non-invasive and objective (albeit expensive) avenue for impairment assessment as certain brain networks signify presence of olfactory dysfunction. Creating a larger training set of patients and controls along with adding additional variables to the CPM may increase its discriminating power. As mentioned earlier, structural differences from voxel-based morphometry have showed many differences between olfactory impairment groups. Thus, combining both structural and functional imaging measures may make the model more specific (e.g., discriminate hyposmia from anosmia).

## Data Availability

Not available

## Supporting Information (SI)

***SI Tables***. Supplementary Table 1 - Statistics of demographic and behavioral variables Supplementary Table 2 - Significant clusters of BOLD response to odor – odorless (regardless of group) Supplementary Table 3 - Statistical values of equivalence tests of parameter estimates from peak voxels extracted from piriform clusters listed in Table 2 between the normosmia and anosmia groups Supplementary Table 4 - Significant clusters of BOLD response to odor – odorless showing linear effect with olfactory function (by group) Supplementary Table 5-Top 10% high-degree nodes obtained from CPM analysis Supplementary Figure 1 - Participant brain trauma severity score and extent score

Supplementary Information appended after references.

## ACKNOWLEDGMENTS

We would like to thank Nicole Reither and Thomas Hummel for assistance with data acquisition and Dustin Scheinost for help in implementing Connectome-based Predictive Modeling. This work was supported by the National Institutes of Health training grant T32NS041228 (M.C.F.); the Yale Medical School Fellowship (M.C.F.). Funding for open access to this research was provided by University of Tennessee’s Open Publishing Support Fund. This publication/paper has been produced benefiting from the 2232 International Fellowship for Outstanding Researchers Program of TÜBİTAK (Project No. 118C299) to M.G.V. However, the entire responsibility of the publication/paper belongs to the owner of the publication/paper. The financial support received from TÜBİTAK does not mean that the content of the publication is approved in a scientific sense by TÜBITAK.

---

**Supplementary Table 1.**
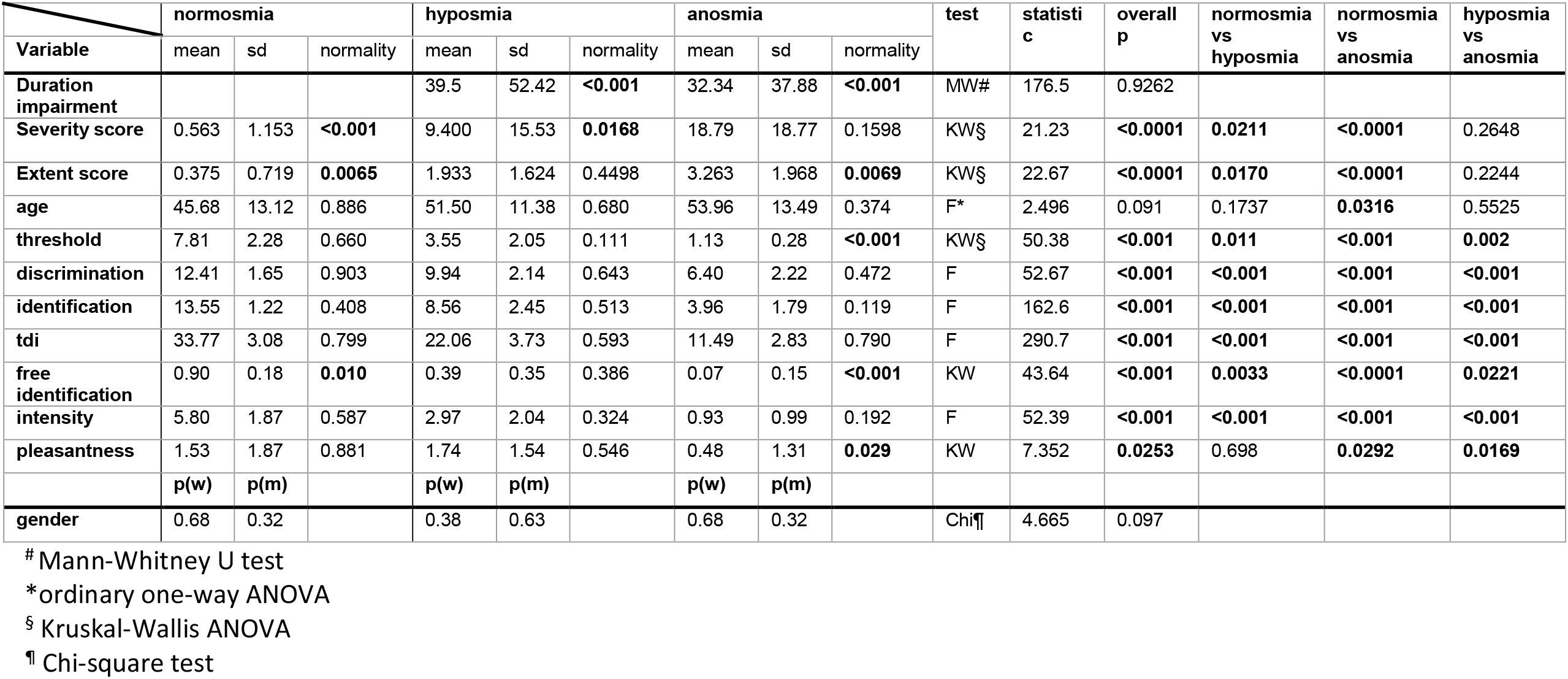
Statistics of demographic and behavioral variables

**Supplementary Table 2.** Significant clusters of BOLD response to odor – odorless (regardless of group)

| Label (Brodmann Area) | P<sub>FWE</sub> | cluster size | T-value | x y z (MNI) |
| --- | --- | --- | --- | --- |
| Intra parietal sulcus | .003* | 243 | 5.620 | -36 -60 54 |
|  |  |  | 3.565 | -50 -60 48 |
|  |  |  | 3.554 | -42 -48 50 |
|  |  |  | 3.338 | -36 -64 40 |
| Cerebellum | .001* | 269 | 4.755 | 22 -76 -22 |
|  |  |  | 4.124 | 36 -74 -28 |
|  |  |  | 3.697 | 46 -68 -28 |
|  |  |  | 3.565 | 20 -86 -20 |
|  |  |  | 3.321 | 24 -68 -28 |
|  |  |  | 3.002 | 24 -88 -12 |
| Inferior frontal gyrus | <.001* | 328 | 4.695 | -54 16 26 |
|  |  |  | 4.069 | -52 12 42 |
|  |  |  | 3.428 | -44 20 30 |
|  |  |  | 3.186 | -54 20 10 |
|  |  |  | 2.788 | -50 8 24 |
| Posterior piriform | ¶ | 44 | 4.053 | -18 -8 -8 |
|  |  |  | 3.226 | -20 -16 -12 |
| Anterior piriform | .007§ | 43 | 4.015 | 26 2 -20 |
| Anterior piriform/ventral insula | ¶ | 26 | 3.564 | -32 8 -16 |
| Posterior piriform | ¶ | 7 | 3.000 | 22 -8 -14 |

§ p-value for alpha = .05 FWE-corrected for multiple comparisons at the voxel level across a small volume search (SVC)
\* p-value for alpha = .05 FWE-corrected for multiple comparisons at the cluster-level across the whole brain
¶ not significant corrected for multiple comparisons

**Supplementary Table 3.**
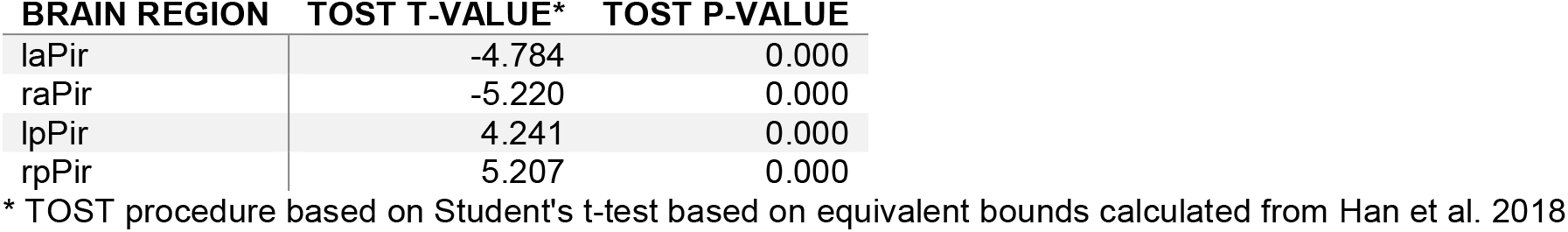
Statistical values of equivalence tests of parameter estimates from peak voxels extracted from piriform clusters listed in Table 2 between the normosmia and anosmia groups

**Supplementary Table 4.** Significant clusters of BOLD response to odor – odorless showing linear effect with olfactory function (by group)

| Label (Brodmann Area) | P<sub>FWE</sub> | CLUSTER SIZE | F-VALUE | X Y Z (MNI) |
| --- | --- | --- | --- | --- |
| Anterior insula | .001§ | 77 | 4.497 | 44 14 6 |
| Posterior cingulate gyrus | .042* | 108 | 4.170 | 0 -22 48 |
|  |  |  | 3.855 | -12 -28 44 |
|  |  |  | 3.365 | 6 -28 48 |
| Ventromedial prefrontal gyrus | <.001* | 324 | 3.995 | 2 46 -6 |
|  |  |  | 3.525 | -2 56 -4 |
|  |  |  | 3.257 | 6 54 10 |
|  |  |  | 3.061 | -2 58 4 |
|  |  |  | 2.934 | 10 44 -2 |
| Anterior insula | .031§ | 50 | 3.619 | -34 16 10 |
| Medio dorsal thalamus | .017§ | 22 | 3.530 | -6 -22 8 |
| Medio dorsal thalamus | .038§ | 22 | 3.252 | 10 -18 10 |

§ p-value for alpha = .05 FWE-corrected for multiple comparisons at the voxel level across a small volume search (SVC)
\* p-value for alpha = .05 FWE-corrected for multiple comparisons at the cluster-level across the whole brain

**Supplementary Table 5.** Top 10% high-degree nodes obtained from CPM analysis

| **NODE** | **DEGREE** | **ANATOMICAL LABEL** | **COORDINATES (MNI)** | **LOBE** | **NETWORK** |
| --- | --- | --- | --- | --- | --- |
| **220** | 7 | Ventral Anterior Cingulate | -3.83,-5.05,32.6 | Limbic | Limbic |
| **218** | 6 | Premotor + Supplementary Motor | -7.75,-22.37,46.05 | Limbic | Motor |
| **39** | 6 | Primary Sensory | 20.01,-33.24,69.77 | Parietal | Motor |
| **260** | 5 | Caudate | -14.6,-3.51,21.09 | Subcortical | Basal Ganglia |
| **216** | 5 | Primary Visual | -22.11,-66.7,7.45 | Occipital | Visual I |
| **197** | 5 | Superior Temporal Gyrus | -57.05,-14.52,-6.87 | Temporal | Motor |
| **188** | 5 | Temporal Pole | -49.73,6.39,-15.15 | Temporal | Motor |
| **132** | 5 | Brainstem | 6.31,-24.92,-17.47 | Brainstem | Cerebellum |
| **15** | 5 | Frontal Eye Fields | 6.69,21.42,31.46 | Prefrontal | Limbic |
| **234** | 4 | Parahippocampus | -30.54,-23.92,-26.61 | Limbic | Basal Ganglia |
| **227** | 4 | AgrRetrolimb? | -7.47,-42.12,13.32 | Limbic | Default Mode |
| **221** | 4 | Ventral Anterior Cingulate | -5.1,13.15,28.74 | Limbic | Limbic |
| **187** | 4 | Temporal Pole | -49.49,11.11,-30.56 | Temporal | Medial Frontal |
| **163** | 4 | Premotor + Supplementary Motor | -56.98,-3.43,6.82 | Motor Strip | Motor |
| **162** | 4 | Premotor + Supplementary Motor | -9.06,0.38,66.53 | MotorStrip | Medial Frontal |
| **160** | 4 | Premotor + Supplementary Motor | -16.2,-19.23,69.53 | MotorStrip | Motor |
| **150** | 4 | Frontal Eye Fields | -5.03,17.67,46.05 | Prefrontal | Medial Frontal |
| **145** | 4 | dIPFC (dorsal) | -10.15,55.69,30.24 | Prefrontal | Medial Frontal |
| **125** | 4 | Putamen | 14.03,8.29,-9.5 | Subcortical | Basal Ganglia |
| **84** | 4 | Ventral Anterior Cingulate | 5.25,-1,35.56 | Limbic | Motor |
| **69** | 4 | Fusiform | 55.22,-56.3,-4.78 | Temporal | Visual Association |
| **44** | 4 | Visual Motor | 7.48,-57.28,61.79 | Parietal | Limbic |
| **42** | 4 | Visual Motor | 14.82,-68.41,34.88 | Parietal | Visual I |
| **34** | 4 | Insula | 41.79,4.97,-7.62 | Insula | Motor |
| **5** | 4 | Ant PFC | 8.18,45.93,-1.72 | Prefrontal | Default Mode |
| **261** | 3 | Putamen | -24.78,5.62,-0.08 | Subcortical | Basal Ganglia |
| **248** | 3 | Cerebellum | -8,-68.4,-19.85 | Cerebellum | Cerebellum |

**Supplementary Figure 1.**
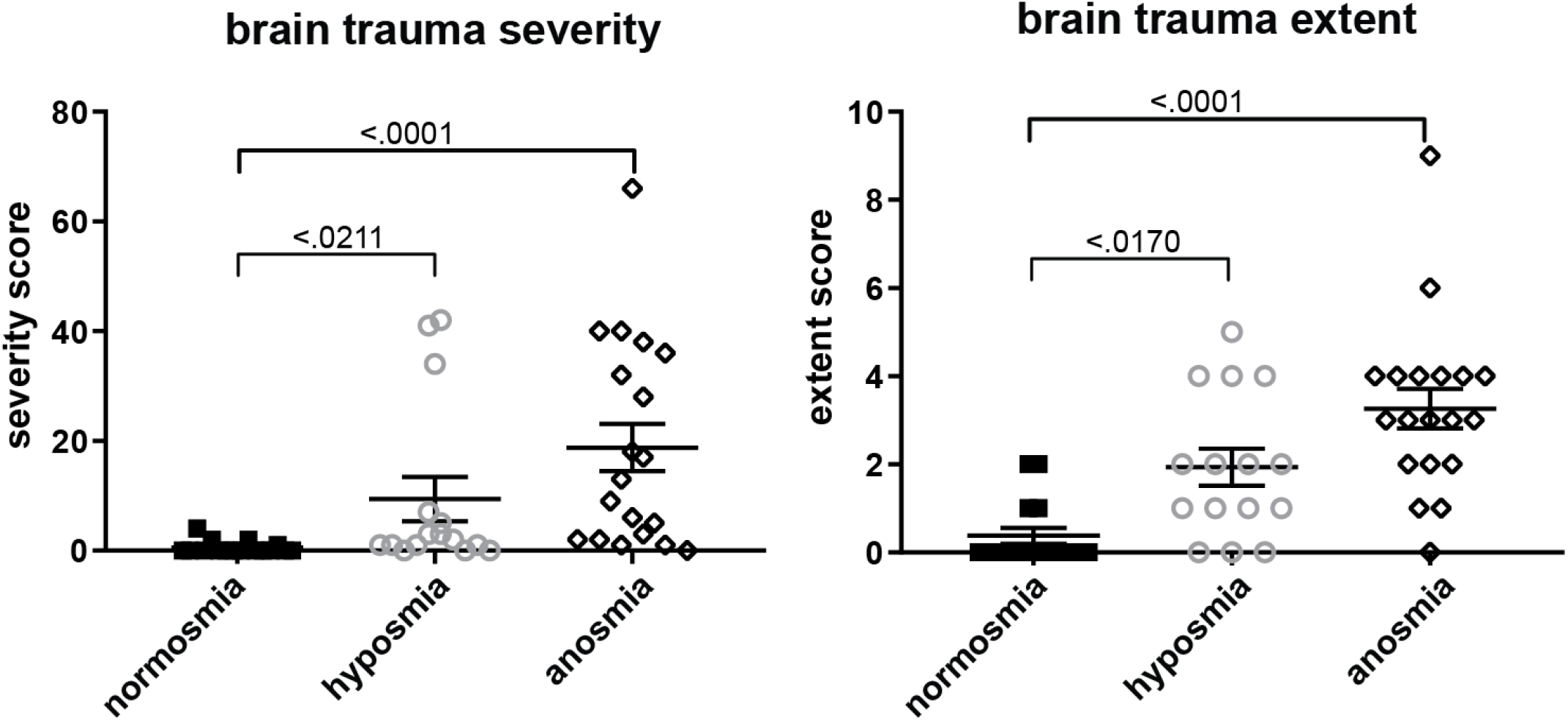
Participant brain trauma severity score and extent score. Left panel shows mean (center line) severity score with standard error of the mean bars (shorter upper and lower bars), with individual data points overlaid for normosmia (solid black squares), hyposmia (open gray circles), and functional anosmia (open black diamonds) participant groups. Severity score reflects the severity of the damage. Right panel shows the mean extent score, with higher numbers indicating more brain regions affected, regardless of how severe the damage was within each brain regions. Bracket indicates significant post-hoc t-test between groups (corrected for multiple comparisons) with p-value.

